# Heterogeneous Associations of Socioeconomic Status with Metabolic Disease in Racial and Ethnic Subgroups in the United States: A Cross-Sectional Cohort Study in NHANES and *All Of Us*

**DOI:** 10.64898/2025.12.08.25341847

**Authors:** Sara J Cromer, Julie E. Gervis, Sherri-Ann M. Burnett-Bowie, Chirag J. Patel

**Affiliations:** Harvard Medical School; Boston, MA; Division of Endocrinology; Massachusetts General Hospital; Boston, MA; Center for Genomic Medicine; Massachusetts General Hospital; Boston, MA; Department of Biomedical Informatics; Harvard Medical School; Boston, MA

## Abstract

**Background:** Unfavorable socioeconomic status (SES) is associated with adverse health outcomes and is believed to at least partially mediate racial and ethnic health disparities such that some authors have recommended incorporation of SES measures in clinical risk algorithms. However, whether the associations of improved SES with improved health are uniform across US racial and ethnic subpopulations and optimal strategies for modeling SES across populations are unknown.

**Methods and Findings:** Among adult participants in the National Health and Nutrition Examination Survey 1999-2018 and the *All of Us* cohort, we used logistic regression to examine the association between SES measures (education and income) with type 2 diabetes (T2D) and obesity prevalence, in the overall population as well as within subgroups of self-reported race and ethnicity. We tested for race or ethnicity-by-SES interactions, modeled SES measures continuously and categorically, and performed mediation analyses to examine the proportion of SES effect on T2D which was mediated by obesity in each racial or ethnic subgroup.

**Results:** Age-adjusted rates of T2D and obesity were highest among non-Hispanic Black, Mexican American, Other Hispanic, and Other/Multi-Racial participants and in those with lower SES. In stratified analyses, higher educational attainment and income were independently associated with lower rates of prevalent T2D and obesity among non-Hispanic White and Asian participants, with smaller or even reversed associations observed among other racial and ethnic groups, particularly non-Hispanic Black participants, in both data sets. Heterogeneity was confirmed by significant race-by-socioeconomic interaction terms. SES measures demonstrated variable patterns of association with disease (e.g., linear, threshold, or U-shaped associations) based on the SES measure, outcome, racial or ethnic group, and data set used. The association between educational attainment and T2D was significantly mediated through BMI among non-Hispanic White participants (32-35% mediation) but not among non-Hispanic Black, Mexican American, and Other Hispanic participants (0-11% mediation).

**Conclusions:** The associations between SES and metabolic disease, the pathways mediating them, and the patterns of association of SES with disease are heterogeneous or even reversed in different US racial and ethnic groups. To prevent biased estimates in both research and clinical calculators which increasingly attempt to incorporate social determinants of health, researchers and clinicians should examine for heterogeneity between groups, particularly racial and ethnic groups.

## INTRODUCTION

Racial and ethnic minoritization and unfavorable socioeconomic status (SES) are associated with numerous adverse health outcomes in the United States (US), including metabolic diseases such as type 2 diabetes (T2D) and obesity.^1–3^ With the growing movement to understand and study race as a social construct with health effects mediated by interpersonal, institutional, and structural racism resulting in inequitable treatment and distribution of resources, many scientists are working to replace race with more proximal and causal mediators of disparities such as SES in health research and even clinical care. However, our understanding of the role of SES, within and between groups in the US, is limited by several factors.

First, an underlying assumption in most studies relating SES with disease outcomes is that the association is homogenous across population subgroups; stated differently, most analyses assume that an achievement of similar SES, for example attainment of a college degree, will confer a similar health benefit in all individuals. However, anecdotal evidence suggests reduced benefits of favorable SES among minoritized populations. For example, in a study of male graduates of Yale University in 1970, Black men had a mortality rate roughly triple that of their White peers by age 60.^4,5^ Similarly, a sociologist noted as early as the 1990s that while there seemed to be racial disparities in health across all socioeconomic strata, “for some indicators of health status…the racial gap becomes larger as SES increases.”^6^ A large study demonstrated variable associations between SES and metabolic disease, namely obesity, have been described between nations at differing stages of economic development,^7^ suggesting that the association of SES with disease is not monolithic and co-varies with other factors in the environment. However,this relationship has been less explored in contemporary subpopulations of the United States, although some studies suggest “diminished returns” of favorable SES in marginalized populations.^8–15^ Thus, the assumption that favorable SES benefits all population groups equally requires further study if we are to understand the intersection of SES and disease.

Second, there is no gold standard by which to measure and analyze SES and its association with disease. SES is a social construct which cannot be directly measured and is therefore frequently proxied by measures such as educational attainment, income, and employment status, either at the level of the individual (measuring personal SES) or over a geographic area (measuring the average SES of the individual’s neighborhood).^16–18^ The choice of which proxy measure or measures to use and how to transform them (e.g., continuous vs. categorized measures of income) varies substantially by study, resulting in limited reproducibility of results related to socioeconomic effects on disease.^19,20^ It is hypothesized the means of transforming variables may lead to different conclusions ultimately and bias research findings, especially when these means are not standardized across studies.^21,22^

In this report, we seek to inform both the intersectionality of race and ethnicity and SES and the impact of SES measure selection and transformation by leveraging ten cycles of the nationally representative National Health and Nutrition Examination Survey (NHANES) and the novel All of Us (AoU) cohort to examine the association of two objective and quantifiable proxy measures of SES – educational attainment and income– with T2D and obesity among individuals of different self-reported racial and ethnic identities. We hypothesized that individuals of different race and ethnicity, who are exposed to different social, cultural, environmental, and structural factors which may impact health, may experience different associations between SES and disease and that the association of SES with disease and the interaction of SES with race and ethnicity will differ based on both the SES measure and its means of transformation.

## METHODS

### Study Population

We leveraged data from (1) the NHANES, a nationally representative sample of community-dwelling individuals in the US, from 1999-2018, and (2) the AoU cohort v7 (workspace identifier: aou-rw-2cd512b1), a national precision medicine initiative enrolling from the general US population, which has generally excellent representation across racial and ethnic groups.^23^ We excluded children (age <18) and those with incomplete information regarding demographics (age, sex, race and ethnicity) or all elements used to define key exposures (education and income), outcomes (prevalent T2D and obesity, see below), or covariates (insurance status, smoking status).

In NHANES, we assigned all participants to a category based on self-reported race and ethnicity, including non-Hispanic White (NHW), non-Hispanic Black (NHB), Mexican American, Other Hispanic, non-Hispanic Asian (NHA), and Other/Multi-Racial race and ethnicity. Because NHA individuals were included among the Other/Multi-Racial category prior to 2011, we separated this category into pre- and post-2011 groups. In AoU, categories based on self-reported race and ethnicity were NHW, NHB, Hispanic, NHA, Multiracial, Other, and None of these.

### Exposure and Outcomes Definitions

The primary exposures were educational attainment and household income-to-poverty ratio (NHANES) or income (AoU). We chose these measures as they can be objectively measured and/or transformed to interpretable units (e.g., years of education, dollars of income) and are among the most commonly used proxy measures of SES.^16,17^ In NHANES, we converted educational attainment from a categorical to a numeric variable, with each 1-unit increase representing approximately a 1-2 year increase in years of education, in order to preserve granularity, allow comparison over a period in which the survey categories were altered, and allow flexibility with modeling (**Supplemental Table 1**). Income-to-poverty ratio was provided within the NHANES and calculated based on annual household income and household size; it had a maximum value of 5.0, with all greater values set to 5.0. In AoU, we converted categorical educational attainment to numeric values matching those assigned in NHANES (**Supplemental Table 1**). In AoU, income was reported as a categorical variable and was transformed to a numeric variable (**Supplemental Table 2**) with each 1 unit increase representing a $25,000 increase in annual income, approximating the federal poverty level for a family of three in 2023.^24^ In both cohorts, a sensitivity analysis was performed categorizing education (no high school (NHANES only), less than high school, high school degree, some college (AoU only), or college degree).

The primary outcomes of this analysis were prevalent T2D and obesity. In NHANES, we defined T2D as the presence of diabetes (any of the following criteria: report of being diagnosed by a healthcare provider with diabetes, use of insulin or other medication for the participant-reported purpose of diabetes or blood sugar control, hemoglobin A1c (HbA1c) ≥ 6.5% (7.8 mmol/L), fasting blood sugar value ≥ 126 mg/dL (7.0 mmol/L), or random blood sugar value ≥ 200 mg/dL (11.1 mmol/L)) without evidence of type 1 diabetes (T1D; which was defined as diagnosis before age 30 years and treatment with insulin only). We defined obesity by the presence of any of the following: report of being told by a healthcare provider that the participant was overweight, use of a prescription medication for the participant-reported purpose of weight control, or body mass index (BMI) ≥ 30 kg/m^2^ among non-NHA participants or ≥ 27.5 kg/m2 among NHA participants.^25^

In AoU, we defined T2D by either the presence of T2D based on self-report, the electronic health record (EHR) diagnoses-based “condition” data set, or maximum HbA1c, fasting glucose, or random glucose consistent with a diagnosis of diabetes based on the criteria listed above; individuals with T1D in the condition data set were excluded. We defined obesity based on either the presence of obesity in the condition data set or a maximum recorded BMI, in either the AoU-collected data or EHR-based data, consistent with obesity based on the criteria listed above. Individuals missing all relevant data (from EHR records and anthropomorphic and blood measures) were excluded from the analysis.

### Associating SES with Outcomes: Statistical Analysis

We conducted all NHANES analyses using weighting procedures to account for complex survey design, as recommended by the NHANES statistical guidance documents.^26^ For baseline characteristics, we reported mean and standard deviation and number and proportion for continuous and categorical measures, respectively. We calculated age-adjusted disease prevalence by standardizing, within each race and ethnicity category, to the projected U.S. population in 2000.^27^

We next performed multivariable logistic regression analyses examining the association between socioeconomic measures and disease outcomes. We performed all analyses initially in the entire sample, then in the entire sample with adjustment for race and ethnicity, and finally in subsamples restricted to single racial and ethnic groups. We adjusted all analyses for key covariates including age, sex, survey year (NHANES only), insurance type and stability (stably insured in the past year with private insurance, stably insured with other insurance, non-stably insured with private insurance (NHANES only), non-stably insured with other insurance (NHANES only), and uninsured; **Supplemental Table 3**), and smoking status (current, former, current or former unspecified (AoU), or never smoker). To test for statistical significance, we performed regression models including race and ethnicity-by-SES interactions to assess for heterogeneity of associations across groups, followed by modified, Satterthwaite-adjusted (NHANES, using survey procedures) and standard likelihood ratio tests examining whether the addition of interaction terms improved model fit.

We performed several sensitivity analyses, including logistic regression analyses using categorized, rather than continuously transformed, SES measures and using alternative model specifications (quasi-binomial, modified Poisson, and Poisson regressions) to confirm the associations between SES and disease outcomes in different racial and ethnic groups.

Following the evaluation for heterogeneity of the effect of SES on disease outcomes across racial and ethnic groups, we performed exploratory analysis examining whether one well-known cause of T2D, elevated BMI, mediated the association between SES and T2D to a similar degree in different groups, using structural equation models ^28^ to quantify the degree to which the association between SES measures and T2D was mediated through BMI.

We performed all analyses using R version 4.0.2 (R Core Team, 2020; Vienna, Austria), including the nhanesA, ggplot2, forestplot, fmsb, metafor, and lmtest packages.^29–33^

### IRB Approval and Role of the Funding Source

The study leveraging de-identified, publicly available data was deemed exempt from institutional review board (IRB) review by the Mass General Brigham IRB.

The funders of the study had no role in study design, data collection, data analysis, data interpretation, or writing of this report.

## RESULTS

### Baseline Characteristics

Between 1999 and 2018, 54,991 adults, representing a weighted population of approximately 217 million community-dwelling civilian adults, participated in the NHANES and had complete covariate data available (**Supplemental Figure 1**). Using weights to account for complex survey design, mean age was 41.2 years, 51.9% were female, and 67.9%, 11.2%, 8.3%, 5.6%, 2.4%, 3.1% and 1.5% identified as NHW, NHB, Mexican American, Other Hispanic, NHA, and Other/Multi-Racial race and ethnicity (pre- and post-2011), respectively (**Table 1**). In AoU, 275,292 had complete covariate data available (**Supplemental Figure 2**). mean age was 51.8 years, 63.4% were female, and 60.3%, 16.1%, 17.1%, 3.2%, 1.7%, 0.6%, and 1.0% identified as NHW, NHB, Hispanic, NHA, Multiracial, Other, and None of these races or ethnicities, respectively (**Table 1**). In both cohorts, NHW and NHA participants were more likely to report higher educational attainment and income-to-poverty ratio (**Supplemental Figure 3**). Educational attainment was modestly correlated with income (Pearson’s coefficient 0.45, p<0.01 in NHANES; 0.46, p<0.01 in AoU). Compared to survey-weighted NHANES, the AoU cohort was older and included more women, fewer individuals who identified as NHW and more who identified as NHB or Hispanic, and more individuals with the highest levels of educational attainment and income.

**Table 1:** Baseline characteristics of participants, including unweighted and weighted sample.

|  | NHANES 2011-2018:<br>Unweighted (n=54,991) | NHANES 2011-2018:<br>Weighted<br>(n=216,871,589) |  | All of Us (n= 275,292) |
| --- | --- | --- | --- | --- |
| Age, mean (SD) | 49.43 (18.66) | 41.77 (13.35) | Age, mean (SD) | 51.81 (16.79) |
| Gender: Female, n (%) | 28,527 (51.9) | 112,606,704 (51.9) | Gender: Female, n (%) | 173720 (63.4) |
| NHANES Survey Year,<br>n (%) |  |  |  |  |
| 1999-2000 | 4,687 (8.5) | 18,026,031 (8.3) |  | - |
| 2001-2002 | 5,240 (9.5) | 20,716,338 (9.6) |  | - |
| 2003-2004 | 4,934 (9.0) | 20,199,151 (9.3) |  | - |
| 2005-2006 | 4,931 (9.0) | 20,913,734 (9.6) |  | - |
| 2007-2008 | 5,884 (10.7) | 21,324,803 (9.8) |  | - |
| 2009-2010 | 6,162 (11.2) | 21,770,096 (10) |  | - |
| 2011-2012 | 5,501 (10.0) | 22,272,803 (10.3) |  | - |
| 2013-2014 | 6,070 (11.0) | 23,609,064 (10.9) |  | - |
| 2015-2016 | 5,865 (10.7) | 23,679,578 (10.9) |  | - |
| 2017-2018 | 5,717 (10.4) | 24,359,993 (11.2) |  | - |
| Race/Ethnicity, n (%) |  |  | Race/Ethnicity, n (%) |  |
| NHW | 24,262 (44.1) | 14,7481,527 (68) | NHW | 165179 (60.3) |
| NHB | 11,474 (20.9) | 24,348,427 (11.2) | NHB | 44149 (16.1) |
| Mexican American | 4,503 (8.2) | 12,136,002 (5.6) |  | - |
| Other Hispanic | 3,004 (5.5) | 5,200,063 (2.4) | Hispanic | 46794 (17.1) |
| NHA | 9,564 (17.4) | 17,819,748 (8.2) | NHA | 8668 (3.2) |
| Other/Multi-Racial<br>including NHA (pre-2011) | 1,331 (2.4) | 6,657,433 (3.1) | Multiracial | 4660 (1.7) |
| Other/Multi-Racial<br>(post-2011) | 853 (1.6) | 3,228,389 (1.5) | Other | 1759 (0.6) |
|  |  |  | None of these | 2779 (1.0) |
| Educational Attainment<br>(Continuous Educational<br>Attainment Value), n<br>(%) |  |  | Educational Attainment<br>(Continuous Educational<br>Attainment Value), n<br>(%) |  |
| Less Than 9th Grade<br>(1) | 6,047 (11.0) | 12,828,321 (5.9) |  | - |
| Less Than High<br>School Degree (2) | 1,070 (1.9) | 2,966,055 (1.4) | Less than a high<br>school degree or<br>equivalent (2) | 21144 (7.7) |
| 9-11th Grade (3) | 7,307 (13.3) | 22,067,379 (10.2) |  | - |
| High School Degree<br>or GED (4) | 11,105 (20.2) | 43,528,777 (20.1) | Highest Grade:<br>Twelve Or GED (4) | 46676 (17.0) |
| High School Degree<br>or GED or Some<br>College or<br>Associates Degree<br>(5) | 3,134 (5.7) | 13,375,693 (6.2) |  | - |
| Some College or<br>Associates Degree<br>(6) | 13,049 (23.7) | 56,262,998 (25.9) | Highest Grade:<br>College One to Three<br>(6) | 69533 (25.4) |
| College Degree or<br>Higher (7) | 11,887 (21.6) | 60,566,209 (27.9) | College graduate or<br>advanced degree (7) | 134832 (49.2) |
| Do not know | 83 (0.2) | 157,130 (0.1) |  | - |
| Missing | 1,295 (2.4) | 5,091,022 (2.3) | PMI: Prefer Not To<br>Answer | 411 (0.2) |
| Refused | 14 (0.0) | 28,005 (0) | PMI: Skip | 1392 (0.5) |
| Continuous Educational<br>Attainment,<sup>a</sup> mean (SD) | 4.70 (1.94) | 5.36 (1.66) | Continuous Educational<br>Attainment,<sup>a</sup> mean (SD) | 5.84 (1.55) |
| Household Income, n<br>(%) |  |  | Household Income, n<br>(%) |  |
|  | - | - | Annual Income: more<br>200k (9) | 21008 (7.7) |
|  | - | - | Annual Income: 150k<br>200k (7.5) | 15005 (5.5) |
| \$100,000 or more | 5,476 (10.0) | 32,324,052 (14.9) | Annual Income: 100k<br>150k (5.5) | 31794 (11.6) |
| \$75,000-99,999 | 3,047 (5.5) | 1,5606,953 (7.2) | Annual Income: 75k<br>100k (4) | 25338 (9.2) |
| \$75,000 or more | 3,714 (6.8) | 20,321,616 (9.4) | | - |
| \$65,000-74,999 | 2,569 (4.7) | 12,621,002 (5.8) | Annual Income: 50k<br>75k (3) | 31553 (11.5) |
| \$55,000-64,999 | 3,184 (5.8) | 14,437,844 (6.7) | | - |
| \$45,000-54,999 | 4,320 (7.9) | 18,357,175 (8.5) | | - |
| \$35,000-44,999 | 5,210 (9.5) | 20,068,404 (9.3) | Annual Income: 35k<br>50k (2) | 22739 (8.3) |
| \$25,000-34,999 | 6,380 (11.6) | 21,752,069 (10) | Annual Income: 25k<br>35k (2) | 19522 (7.1) |
| \$20,000-24,999 | 4,146 (7.5) | 12,934,326 (6) | | - |
| Over \$20,000 | 1,608 (2.9) | 5,882,085 (2.7) | | - |
| Under \$20,000 | 443 (0.8) | 1,054,752 (0.5) | | - |
| \$15,000-19,999 | 3,875 (7.0) | 11,167,767 (5.1) | Annual Income: 10k<br>25k (1) | 29982 (10.9) |
| \$10,000-14,999 | 3,988 (7.3) | 10,684,859 (4.9) | Annual Income: less<br>10k (1) | 31977 (11.7) |
| \$5,000-9,999 | 2,485 (4.5) | 6,516,581 (3) | | - |
| \$0-4,999 | 1,157 (2.1) | 3,193,678 (1.5) | PMI: Prefer Not To<br>Answer | 32517 (11.9) |
| Missing | 3,389 (6.2) | 9,948,425 (4.6) | PMI: Skip | 12553 (4.6) |
| Income-to-Poverty<br>Ratio, mean (SD) | 2.52 (1.62) | 3.28 (1.58) | Income (per \$25,000),<br>mean (SD) | 3.58 (2.56) |
| Insurance Type and<br>Stability,<sup>b</sup> n (%) |  |  | Insurance Type and<br>Stability,<sup>b</sup> n (%) |  |
| Stably Insured –<br>Private | 27,346 (49.7) | 128,978,929 (59.5) | Stably Insured – Private | 143754 (52.5) |
| Stably Insured –<br>Other | 13,198 (24.0) | 38,271,607 (17.6) | Stably Insured – Other | 114976 (42.0) |
| Unstably Insured –<br>Private | 1,798 (3.3) | 7,732,641 (3.6) |  | - |
| Unstably Insured –<br>Other | 1,336 (2.4) | 3,789,919 (1.7) |  | - |
| Uninsured | 11,313 (20.6) | 38,098,492 (17.6) | Uninsured | 15258 (5.6) |

<sup>a</sup> See Supplemental Table 1
<sup>b</sup> See Supplemental Table 2
NHANES = National Health and Nutrition Examination Survey; NHW = non-Hispanic White; NHB = non-Hispanic Black; NHA = non-Hispanic Asian.

### Association Between SES and T2D and Obesity Prevalence

In both cohorts, age-standardized prevalence of T2D was lower among NHW participants than among participants of other race and ethnicity (**Figure 1**). For example, NHB participants with a college degree had similar (NHANES) or even higher (AoU) T2D prevalence compared to NHW individuals with only some high school education (less than a high school degree). Similarly, age-standardized prevalence of obesity varied between racial and ethnic groups. NHB, Mexican American, and other Hispanic participants (NHANES) and NHB participants (AoU) with college degrees had higher rates of obesity than NHW individuals of any educational attainment, including those with less than a 9^th^ grade education.

**Figure 1:**
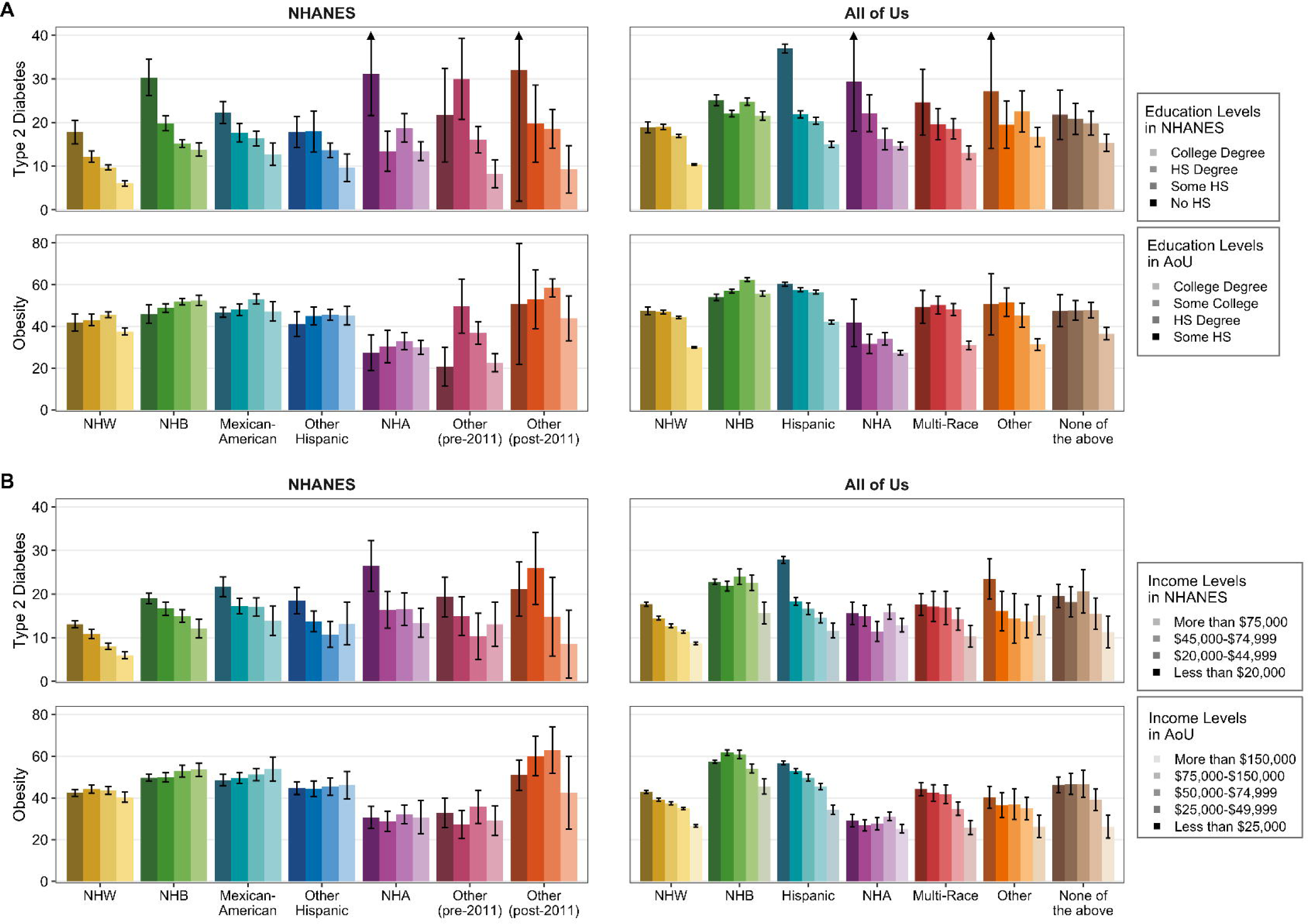
Age-standardized type 2 diabetes and obesity prevalence by race and ethnicity and by educational attainment (A) or income (B) in NHANES (left column) and AoU (right column).

In adjusted models, higher educational attainment was associated with lower prevalence of T2D in the entire population and in each race and ethnicity group except NHA (NHANES) or Other race or ethnicity (AoU; **Figure 2A**; **Supplemental Table 4; Supplemental Figure 4A**). However, the effect size varied by race or ethnicity group. For instance, each 1 unit increase in continuous education measure (equivalent to approximately 1-2 years increase in educational attainment) was associated with a 12% decrease in T2D prevalence among NHW participants (OR 0.88 (95% CI 0.85, 0.91)) but a 4% decrease in prevalence among NHB participants (OR 0.96 (95% CI 0.92, 0.99)) in NHANES. In AoU, a similar discordance occurred with NHW participants experiencing a 19% decrease in T2D prevalence (OR 0.81 (95% CI 0.80, 0.82)) compared to only a 4% decrease in NHB participants (OR 0.96 (95% CI 0.94, 0.97)). Interaction analyses demonstrated significant race-by-SES interactions for NHB and Mexican-American individuals in NHANES and for all non-NHW racial and ethnic groups in AoU (**Table 2**). Modified (NHANES) and standard (AoU) likelihood ratio tests demonstrated that the addition of race-by-SES interactions significantly improved model fit in both cohorts (**Supplemental Table 5**).

**Figure 2:**
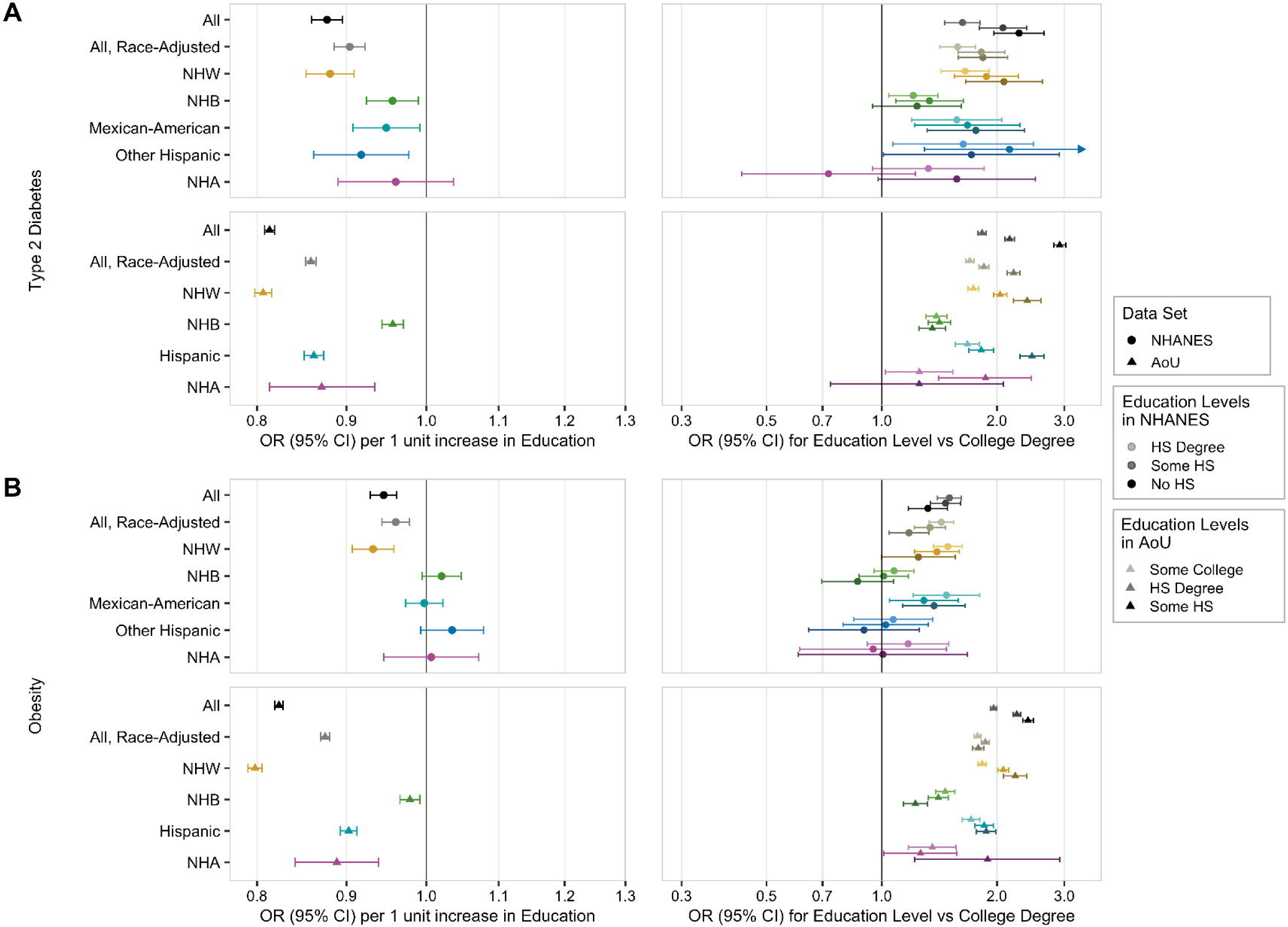
Forest plots of the adjusted association of educational attainment, modeled continuously (left column) or categorically (right column), with (A) type 2 diabetes and (B) obesity prevalence, in the population overall and in each racial and ethnic subgroup, in NHANES (circles) and AoU (triangles).

**Table 2:**
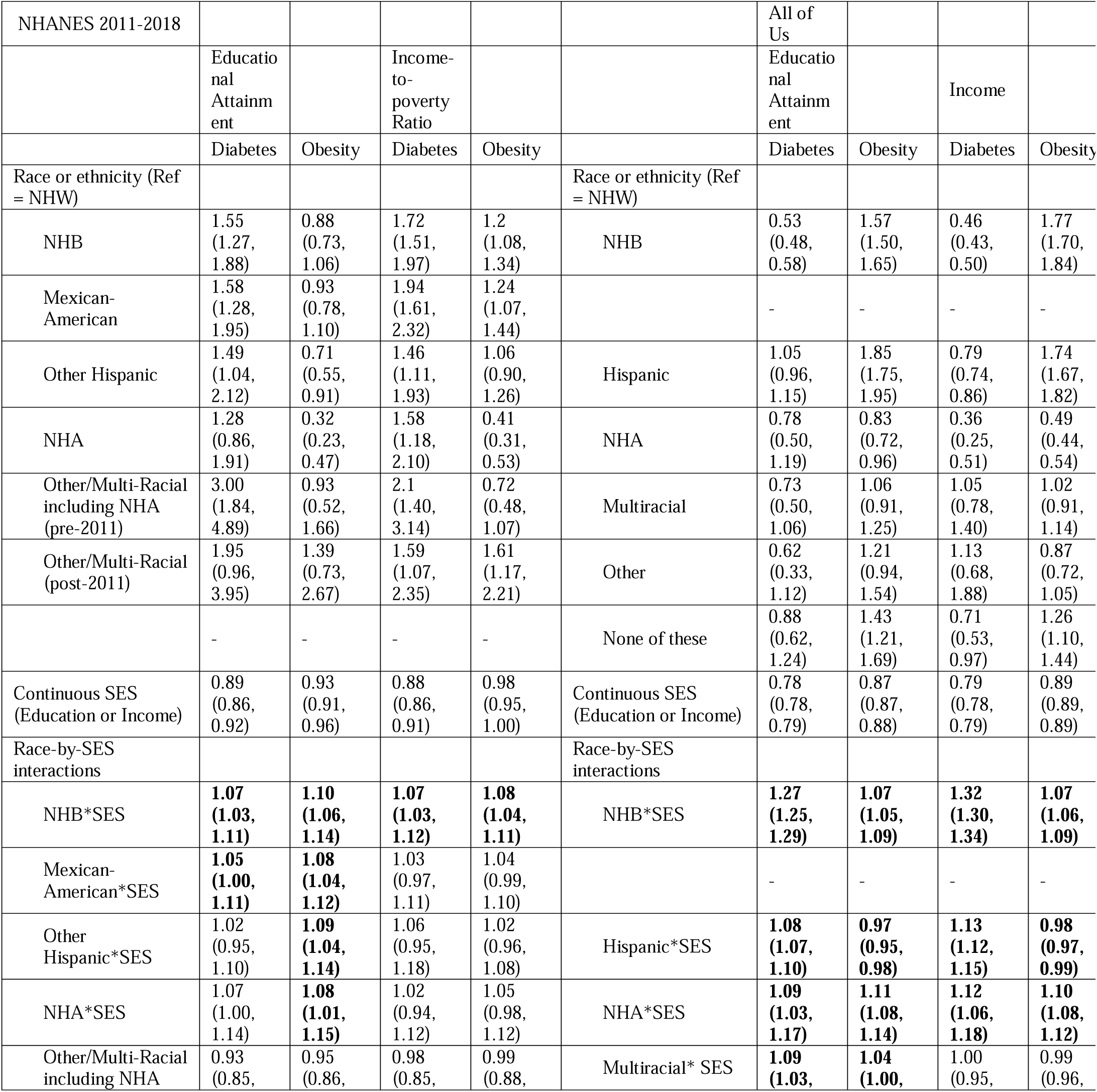
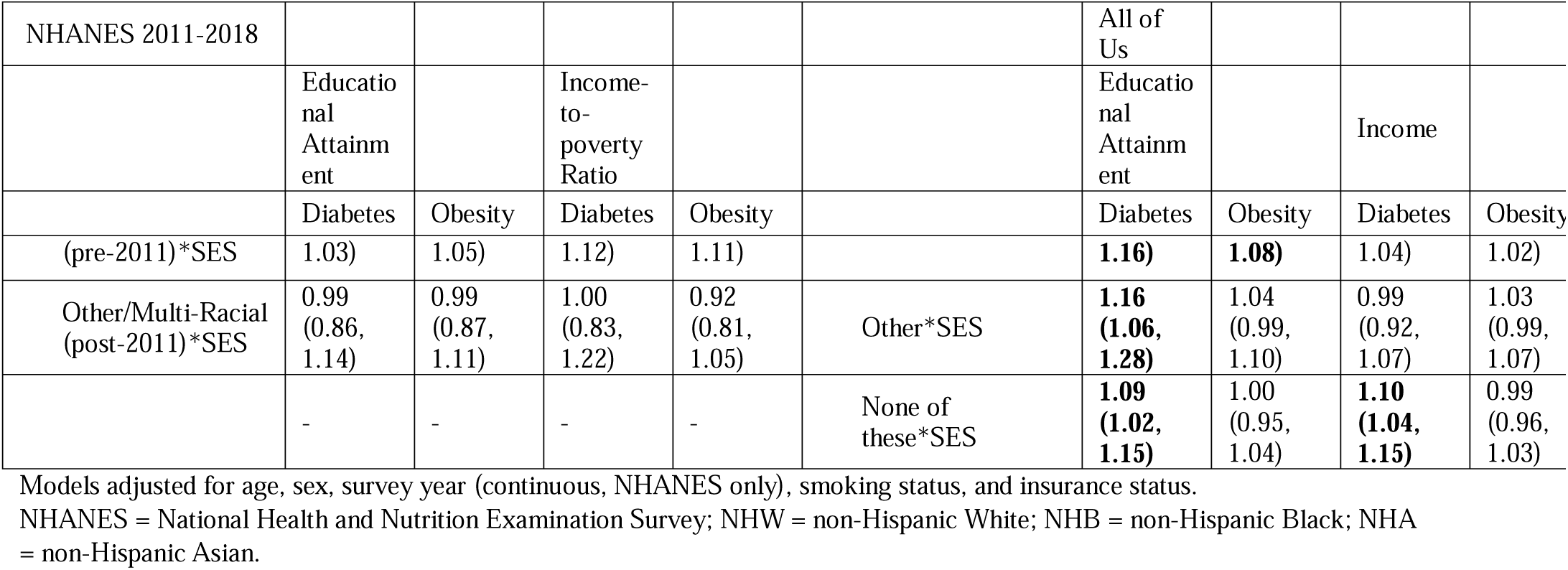
Interaction between race or ethnicity and socioeconomic status in fully adjusted models.

By contrast, the relationship between higher educational attainment and obesity was significant among NHW participants (OR 0.93 (95% CI 0.91, 0.96)) but not significant within any other racial or ethnic group in NHANES. The relationship between education and obesity in NHW participants in AoU was also significantly attenuated in almost all other groups in AoU (**Figure 2B; Supplemental Table 4; Supplemental Figure 4B**). Interaction analyses demonstrated significant race-by-SES interactions for NHB, Mexican-American, Other Hispanic, and NHA individuals in NHANES and for NHB, Hispanic, NHA, and Multiracial individuals in AoU (**Table 2**). Likelihood ratio tests demonstrated that the addition of race-by-SES interactions significantly improved model fit in both cohorts (**Supplemental Table 5**).

Differences in the association between income and T2D prevalence in racial and ethnic subgroups were less pronounced with overlapping confidence intervals than observed between educational attainment and T2D, although the OR remained largest among NHW individuals in both data sets (**Figure 3A; Supplemental Table 4; Supplemental Figure 5A**) with one exception: NHA participants in AoU experienced a smaller reduction in T2D prevalence compared to other participants (e.g., OR 0.96 (95% CI: 0.93, 0.99) for NHA v. OR 0.85 (95% CI: 0.85, 0.86) for NHW). Interaction analyses demonstrated significant race-by-SES interactions for NHB individuals in NHANES and for NHB, Hispanic, NHA, and None of these racial and ethnic groups in AoU (**Table 2**). Likelihood ratio tests demonstrated that addition the of race-by-SES interactions significantly improved model fit in AoU but not in NHANES (**Supplemental Table 5**).

**Figure 3.**
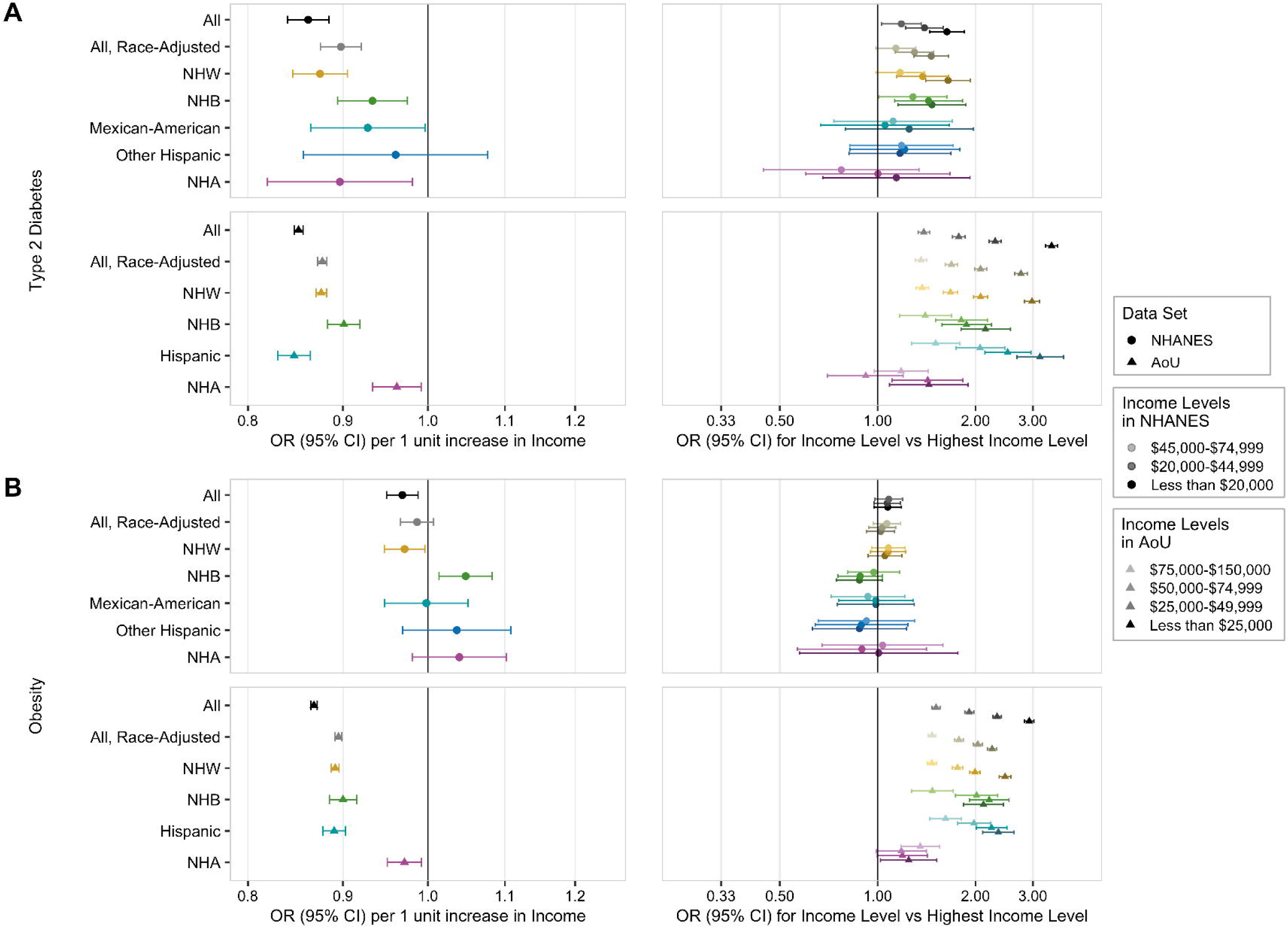
Forest plots of the adjusted association of income, modeled continuously (left column) or categorically (right column), with (A) type 2 diabetes and (B) obesity prevalence, in the population overall and in each racial and ethnic subgroup, in NHANES (circles) and AoU (triangles).

The racial and ethnic difference in effect size was perhaps most striking for the relationship between income and obesity in NHANES (**Figure 3B**; **Supplemental Table 4; Supplemental Figure 5B**), with significant protective associations in the NHW cohort (OR 0.97 (95% CI 0.95, 0.996) not only attenuating but even reversing in the NHB cohort (OR 1.05 (95% CI 1.01, 1.08). We found no significant association for other racial and ethnic groups (Figure 2B) in NHANES. In AoU, this reversal of association was not observed; however, NHA participants had a significantly less protective association between income and obesity than other groups (OR 0.97 (95% CI: 0.95, 0.99) for NHA v. OR 0.89 (95% CI: 0.89, 0.90) for NHW; **Figure 3B**). Interaction analyses demonstrated significant race-by-SES interactions for NHB individuals in NHANES and for NHB, Hispanic, and NHA individuals in AoU (**Table 2**). Likelihood ratio tests demonstrated that the addition of race-by-SES interactions significantly improved model fit in both cohorts (**Supplemental Table 5**).

Sensitivity analyses examining categorized educational attainment and income demonstrated similar results, with greater protective effects of higher education and income among NHW individuals than among NHB individuals (**Figures 2,3**). However, categorical analyses demonstrated that modeling of either SES measure as a continuous value with a linear association with either outcome may not be appropriate across diverse racial and ethnic groups. For example, in AoU, decreasing categories of educational attainment were associated with a step-wise increase in risk for T2D in the overall cohort and in NHW individuals, but threshold effects existed in other racial and ethnic groups – anything less than a college degree was associated with approximately 50% increased odds of T2D among NHB individuals, with no differences between lesser levels of education, while having either a high school degree or at least some high school education resulted in approximately 80% increased odds of T2D for Hispanic individuals, compared to having a college degree, but having no high school education at all resulted in approximately 150% increased odds of T2D (**Figure 2A**). For other exposure-outcome pairs, their seemed to be U- or J-shaped curves, as well. For example, in AoU, NHW individuals with some college education seemed to have higher risk for T2D compared to NHW individuals with either higher or lower educational attainment. Similarly, NHB individuals with some college education seemed to have higher risk for obesity than NHB individuals with either higher or lower educational attainment (**Figure 2B**).

Sensitivity analyses examining alternative modeling strategies (quasi-binomial, modified Poisson, and Poisson regressions) were consistent with the primary results (**Supplemental Figures 6,7**).

### Exploratory Analysis: Mediation of the Association between SES and T2D by BMI

In addition to exploring heterogeneity of the association between SES and T2D across racial and ethnic groups, we aimed to explore whether this association was similarly mediated by differences in BMI between groups. We examined the proportion of the effect of socioeconomic measures on T2D prevalence which was mediated by differences in BMI, overall and within each population subgroup. We found the proportion of this association mediated through BMI varied by racial or ethnic subgroup. For example, 34.5% of the association between educational attainment and T2D prevalence was mediated through BMI among NHW participants in NHANES, while only 5-8% of this association was mediated through BMI among NHB, Mexican American, and Other Hispanic participants (**Table 3**). Similarly, 32.3% of the association between educational attainment and T2D prevalence was mediated through BMI among NHW participants in AoU, while only 1.3% and 11.2% of this association mediated through BMI among NHB and Hispanic participants in AoU. While 22.6% of the association between income-to-poverty ratio and T2D prevalence was mediated through BMI among NHW participants in NHANES, only 0-16% of this association was mediated through BMI among NHB and Mexican American participants (**Table 3**).

**Table 3:** Total effect, direct effect, and indirect effect (through body mass index) of educational attainment and income on type 2 diabetes prevalence and proportion mediated through body mass index, within the overall population and distinct racial and ethnic subpopulations.

|  | Educational Attainment |  |  |  | Income |  |  |  |
| --- | --- | --- | --- | --- | --- | --- | --- | --- |
| Race and Ethnicity Group | Total Effect | Direct Effect | Indirect Effect | Proportion Mediated | Total Effect | Direct Effect | Indirect Effect | Proportion Mediated |
| NHANES |  |  |  |  |  |  |  |  |
| All | -0.146 | -0.112 | -0.034 | 0.234 | -0.15 | -0.123 | -0.027 | 0.18 |
| All-Race-adjusted | -0.112 | -0.084 | -0.028 | **0.315** | -0.105 | -0.085 | -0.019 | **0.26** |
| NHW | -0.137 | -0.09 | -0.047 | **0.345** | -0.123 | -0.096 | -0.028 | **0.22** |
| NHB | -0.044 | -0.042 | -0.002 | **0.051** | -0.066 | -0.066 | 0 | **-0.00** |
| Mexican American | -0.069 | -0.063 | -0.005 | **0.078** | -0.087 | -0.081 | -0.005 | **0.06** |
| Other Hispanic | -0.101 | -0.094 | -0.006 | **0.064** | -0.056 | -0.047 | -0.009 | **0.15** |
| NHA | -0.037 | -0.031 | -0.007 | **0.177** | -0.105 | -0.113 | 0.008 | **-0.07** |
| Other (pre-2011) | -0.174 | -0.149 | -0.025 | **0.145** | -0.101 | -0.091 | -0.01 | **0.09** |
| Other (post-2011) | -0.137 | -0.118 | -0.019 | **0.139** | -0.095 | -0.072 | -0.023 | **0.24** |
| AoU |  |  |  |  |  |  |  |  |
| All | -0.207 | -0.161 | -0.046 | **0.224** | -0.152 | -0.111 | -0.041 | **0.27** |
| All-Race-adjusted | -0.151 | -0.121 | -0.03 | **0.383** | -0.122 | -0.09 | -0.032 | **0.45** |
| NHW | -0.217 | -0.147 | -0.07 | **0.323** | -0.123 | -0.084 | -0.038 | **0.31** |
| NHB | -0.04 | -0.039 | -0.001 | **0.013** | -0.096 | -0.072 | -0.024 | **0.24** |
| Hispanic | -0.145 | -0.129 | -0.016 | **0.112** | -0.156 | -0.133 | -0.022 | **0.14** |
| NHA | -0.12 | -0.08 | -0.04 | **0.334** | -0.025 | -0.016 | -0.009 | **0.35** |
| Multiracial | -0.152 | -0.102 | -0.05 | **0.327** | -0.107 | -0.077 | -0.03 | **0.27** |
| Other | -0.063 | 0.021 | -0.084 | **1.331<sup>a</sup>** | -0.069 | -0.041 | -0.029 | **0.41** |
| None of these | -0.158 | -0.129 | -0.029 | **0.183** | -0.144 | -0.11 | -0.034 | **0.23** |

<sup>a</sup> Negative proportion mediated and proportion mediated >1 should be interpreted as the absence of mediation; both occur in the setting where direct and indirect effects (via the mediator, BMI) of the exposure (SES) on the outcome (type 2 diabetes) are in different directions.
NHANES = National Health and Nutrition Survey; AoU = All of Us cohort; NHW = non-Hispanic White; NHB = non-Hispanic Black. NHA = non-Hispanic Asian; BMI = body mass index.

## DISCUSSION

In this analysis examining the association between SES measures and metabolic disease in a nationally representative US population as well as a large national biobank, both overall and within distinct racial and ethnic groups, we found that the association between SES and disease varies in both magnitude and direction depending on the SES measure used (e.g., education or income), the means of transformation of the SES measure, the disease studied, and the population characteristics of the sample. Specifically, for example, we found that each 1-2 year increase in educational attainment was associated with as little as 4% and as much as 12-19% reduced odds of T2D, depending on racial or ethnic group. For each 1-unit increase in income-to-poverty ratio, NHW individuals experienced 3% reduced odds of obesity while NHB individuals experienced 5% increased odds of obesity. Meanwhile, when SES measures were analyzed categorically, different patterns of association (linear, U-shaped, and threshold effects) were observed in different groups and for different diseases. Our findings suggest that monolithic analyses of the association between SES and disease within the US population may obscure variability which would be observed if examining associations within individual subpopulations, thus leading to an incomplete understanding of the populations at highest risk for disease as well as the potential impact of public health interventions.

A thorough understanding of the interplay of these two risk factors and the potential bias caused by analyzing socioeconomic measures homogeneously across populations is especially important as incorporation of SES in clinical prediction algorithms is already underway.^34,35^ Further, some follow-up studies examining the inclusion of additional individual- and area-level SES measures in clinical algorithms have found statistically significant differences in the association of SES with disease, as well as differences in tool performance across racial and ethnic groups following the addition of these measures.^36^ While efforts to move past “race-based medicine,” further incorporate the most proximal risk factors of disease, and capture sociocultural factors leading to disease are laudatory, we believe greater understanding of the role of SES is needed for its appropriate use in clinical care.

Most analyses examining socioeconomic health disparities examine diverse populations with a few models, reporting a single, non-stratified estimate across all groups. Although this may provide an overall, weighted average of the effect in the general population, our results suggest that it may obscure disparate associations which exist within subpopulations. As most US-based samples include a majority of NHW individuals, the overall result will be weighted to reflect predominantly the association observed within that population. As our analyses suggest that the association between SES and disease among NHW individuals is significantly different from that of other populations, this will systematically obscure important findings among minoritized groups. When reporting associations between SES and disease, future studies should examine for effect modification by race and ethnicity. Further research is also needed to examine whether the association of SES and disease is significantly modified by other factors, such as biological sex or gender, region, physical environment, or year.^37,38^ SES may also impact diabetes risk differently in different population groups. In exploratory analyses notably limited in their interpretation by cross-sectional data collection, we observed that the proportion of the association of SES with T2D prevalence mediated through BMI varied substantially by race and ethnicity, highlighting differences in diabetes pathogenesis, with higher diabetes risk observed at lower BMI for non-White individuals, both in the US and across the world.^39–41^

Limited studies have examined the intersection of race and ethnicity and SES on diabetes outcomes, but these have generally reported similar findings to our study, with SES having different associations with disease in different racial and ethnic groups. For example, a 2014 analysis using the National Health Information Survey examined the prevalence of T2D among poor and nonpoor Black and White individuals living in poor and non-poor neighborhoods. The analysis found that a poor White person living in a poor neighborhood (highest risk group) had more than double the risk of diabetes compared to a non-poor White person living in a non-poor neighborhood (lowest risk group), while the analogous comparison among Black individuals (high individual and neighborhood risk v. low individual and neighborhood risk) was associated with only 17% increased risk of diabetes.^42^ Similar to our findings, this suggests a different and greater impact of favorable SES among White individuals compared to Black individuals.

A larger body of work has examined the intersection of race and ethnicity and SES on obesity outcomes, demonstrating variable associations between SES and obesity rates in countries at different stages of the “obesity transition.”^7^ For countries at earlier stages of this transition, generally lower-income and less industrialized countries, higher SES is associated with improved food security, more sedentary lifestyles, and higher rates of obesity.^43^ However, countries may progress to later stages of the transition in which calorie-dense and sedentary environments affect the entire population, with those of higher SES being able to afford healthier foods and having the resources and time to devote to physical activity, thus leading to lower rates of obesity in high SES groups. At the individual level, some authors in the field have articulated a theory of “diminished returns” of improved SES among marginalized and immigrant populations ^44^. For example, intersectionality of race or ethnicity and SES, has been reported for childhood BMI^10,11^ asthma,^45^ heart disease,^12^ and overall comorbidity burden,^13–15^ with diminished benefits seen in marginalized groups (racially and ethnically minoritized groups, the LGBTQIA community, and American immigrants) in a variety of cross-sectional and cohort studies. For example, a study performed among individuals with osteoarthritis found that higher income was associated with reduced odds of being overweight or obese among White individuals but not among Black individuals,^8^ with a related study showing a similar interaction between educational attainment and sexual orientation on obesity.^9^ This work provides a foundation through which the findings of our study using large, nationally representative data sets may be interpreted.

Our study supports prior evidence documenting marginalization-related diminished returns and suggests that different population groups within a single country may be at different stages of the “obesity transition.” We found that higher SES is associated with lower rates of obesity in NHW Americans but higher rates of obesity among NHB Americans, reflecting associations seen in countries at different stages of this phenomenon, due to different structural pressures influencing the environment, lifestyle, and disease risk of affected individuals. Within individual US states, heterogeneity in the association of individual- and area-level SES with BMI and mortality were also found in different racial and ethnic subgroups in the 1990s to early 2000s, further supporting that this same “obesity transition” phenomenon may exist within local populations.^46–48^ Despite these earlier findings, only a minority of subsequent studies on the association of SES with metabolic disease examined the interaction between SES and other factors, with most studies continuing to analyze SES monolithically across populations.^18,49,50^

Our findings also illustrate that different measures of SES – education and income – have different associations with disease outcomes, suggesting that the choice of socioeconomic measure should be carefully considered, and if possible, standardized across research studies to improve interpretability and reproducibility of results. The use of more than one socioeconomic measure, when available, and carefully planned sensitivity analyses to examine the impact of variable transformation and modeling on study results may better capture diverse elements of SES, improve rigor and reproducibility of SES-related analysis, and have greater predictive value for health outcomes, as seen in previous studies.^51,52^

Finally, it is important to highlight that racial and ethnic disparities in T2D and obesity prevalence far outweigh socioeconomic disparities in our study, with NHB individuals with college degrees having a similar or greater age-standardized prevalence of T2D as NHW individuals with less than a high school degree. Previous studies have similarly documented that SES only partially mediates the association between race and ethnicity and health outcomes.^53,54^ Although efforts to improve socioeconomic barriers to care are critical, they are not sufficient to overcome racial and ethnic disparities in the United States which arise from manifold factors in the environment – including direct and indirect effects of structural racism affecting individual’s physical environments, chronic stress, and access to healthcare and required elements of a healthy lifestyle – that are not captured by simple socioeconomic adjustment.

This study has numerous strengths including large sample size in a nationally representative population. However, our findings must be interpreted considering the study design and data sources. NHANES is a cross-sectional survey, and direct and mediated associations of prevalent factors measured at a single time point may not be causal; however, the age at which participants attain their SES, especially educational attainment, is most likely to come before T2D onset and study entry, making reverse causality unlikely. The categories of race and ethnicity used in NHANES changed in 2011-2012 such that NHA individuals were included with the “Other/Multi-Racial” category before that time but not subsequently; this may lead to diverse populations included within the pre-2011 Other/Multi-Racial category and decreased sensitivity of obesity designations, as the NHA-specific cutoff was applied only to those self-reporting as NHA, a category which only existed beginning in 2011-2012. The conversion of educational attainment from a categorical to a numeric variable assumes a linear relationship between educational attainment and health outcomes and approximately equal distance or effect between adjacent categories of educational attainment; we performed sensitivity analyses using categorical education and income to confirm our results. In NHANES, the income-to-poverty measures have a maximum value of 5.0, treating all participants with the highest income levels as equal. Mediation analyses rely on multiple strong assumptions, particularly that all potential confounding between the exposure, mediator, and outcome has been accounted for, and we cannot be sure that these assumptions are met in this analysis; results of the mediation analysis should be interpreted cautiously. Although the findings in NHANES and AoU were overall consistent, showing heterogeneity in the association between SES and metabolic disease, with NHW individuals generally receiving the greatest relative benefit from increased SES, differences observed between the two analyses may be due to a number of factors including temporal differences in recruitment of participants, minor differences in collection and transformation of the key SES exposure variables, and selection bias in AoU, particularly in the NHA cohort which had very high levels of educational attainment and income. Lastly, our analyses are not adjusted for multiple testing.

In conclusion, we found that the association between measures of SES and T2D and obesity outcomes vary in both magnitude and direction between racial and ethnic subgroups in the United States over the past two decades and currently. Similarly, choice of SES measure and transformation and modeling strategies revealed small but meaningful differences in described associations. In studies examining socioeconomic effects on disease outcomes, associations between SES and clinical outcomes should not be analyzed monolithically nor assumed to be consistent across subpopulations living in different cultural contexts. We recommend stratified analyses, with robust sensitivity analyses examining the effects of SES modeling choices on outcomes, to document heterogeneity between groups and improve the reproducibility of SES-related health research.

## Acknowledgements

We gratefully acknowledge NHANES and *All of Us* participants, without whom this research would not have been possible, for their participation in research. We also thank the National Institutes of Health’s *All of Us* Research Program for making available the data examined in this study.

## Author contributions

SJC conceived of the study, obtained data, performed the analysis, and drafted the manuscript, with expert guidance from CJP. All authors contributed to the interpretation of study results and critical review of the manuscript.

## Data availability statement

NHANES data is publicly available from the CDC website (https://www.cdc.gov/nchs/nhanes/index.htm) and should be handled using best practices outlined by NCHS. All of Us controlled-tier data is available by application to the All of Us Researcher Workbench (workbench.researchallofus.org).

