## Supplemental for "Heterogeneous Associations of Socioeconomic Status with Metabolic Disease in Racial and Ethnic Subgroups in the United States: A Cross-Sectional Cohort Study in NHANES and *All Of Us*"

**Supplemental Appendix**

**Supplemental Table 1: Conversion of categorical educational attainment to a continuous variable.**

|  |  | NHANES |  |  | AoU |  |
| --- | --- | --- | --- | --- | --- | --- |
| Continuous Educational Attainment Value | Categorical Educational Attainment Level | NHANES Educational Attainment Category | NHANES Survey Years Used | Approximate Years of Education | AoU Educational Attainment Category | Approximate Years of Education |
| 1 | No High School | Less Than 9th Grade | 1999-2016 | 0-8 |  |  |
| 2 | Less than High School | Less Than High School Degree | 2017-2018 | 0-11 | Less than High School | 0-11 |
| 3 | Less than High School | 9-11th Grade | 1999-2016 | 9-11 |  |  |
| 4 | High School Degree | High School Degree or GED | 1999-2016 | 12 | High School Degree | 12 |
| 5 | High School Degree | High School Degree or GED or Some College or Associate Degree | 2017-2018 | 12-15 |  |  |
| 6 | High School Degree | Some College or Associate Degree | 1999-2016 | 13-15 | Some College | 13-15 |
| 7 | College Degree or Higher | College Degree or Higher | 1999-2018 | 16+ | College Degree or Higher | 16+ |

**Supplemental Table 2: Conversion of categorical income to a continuous variable (AoU only).**

| Continuous Income Value | AoU Income Categories | Multiples of $25,000 (FPL for family of 3 in 2023) |
| --- | --- | --- |
| 1 | $0-9,999  $10,000-24,999 | 0-1 |
| 2 | $25,000-34,999  $35,000-49,999 | 1-2 |
| 3 | $50,000-74,999 | 2-3 |
| 4 | $75,000-99,999 | 3-4 |
| 5.5 | $100,000-149,999 | 4-6 |
| 7.5 | $150,000-199,999 | 6-8 |
| 9 | $200,000 or more | 8+ |

**Supplemental Table 3: Insurance type and stability variable classification.**

| Insured Currently | Insurance Type | Uninsured in Past Year (NHANES only) | NHANES Insurance Type and Stability Variable Designation | AoU Insurance Type and Stability Variable Designation |
| --- | --- | --- | --- | --- |
| Yes | Private | Yes | Stably Insured-Private | Stably Insured-Private |
| Yes | Other^a^ | Yes | Stably Insured-Other | Stably Insured-Other |
| Yes | Private | No | Unstably Insured-Private | - |
| Yes | Other^a^ | No | Unstably Insured-Other | - |
| No | - | - | Uninsured | Uninsured |

^a^ Other insurance types include: Medicare, Medi-Gap, Medicaid, SCHIP, military, Indian Health Service, state-sponsored, other government-sponsored, and single service plans. If a participant reported both private and other types of insurance, they were coded as privately insured.

**Supplemental Table 4: Adjusted association of educational attainment and income with type 2 diabetes and obesity prevalence, overall and in strata of race and ethnicity (depicted in Figure 2), per 1 unit increase in continuous educational achievement of income (see Supplemental Tables 1-2).**

|  | SES Measure: Educational Attainment | SES Measure: Educational Attainment | SES Measure: Income-to-Poverty Ratio | SES Measure: Income |
| --- | --- | --- | --- | --- |
| **NHANES** | Outcome: T2D | Outcome: Obesity | Outcome: T2D | Outcome: Obesity |
| All | 0.88 (0.86, 0.90) | 0.95 (0.93, 0.96) | 0.86 (0.84, 0.88) | 0.97 (0.95, 0.99) |
| All-Race-adjusted | 0.90 (0.89, 0.92) | 0.96 (0.94, 0.98) | 0.90 (0.88, 0.92) | 0.99 (0.97, 1.01) |
| NHW | 0.88 (0.85, 0.91) | 0.93 (0.91, 0.96) | 0.87 (0.85, 0.90) | 0.97 (0.95, 1.00) |
| NHB | 0.96 (0.92, 0.99) | 1.02 (0.99, 1.05) | 0.93 (0.89, 0.97) | 1.05 (1.01, 1.08) |
| Mexican American | 0.95 (0.91, 0.99) | 1.00 (0.97, 1.02) | 0.93 (0.86, 1.00) | 1.00 (0.95, 1.05) |
| Other Hispanic | 0.92 (0.86, 0.98) | 1.03 (0.99, 1.08) | 0.96 (0.86, 1.08) | 1.04 (0.97, 1.11) |
| NHA | 0.96 (0.89, 1.04) | 1.01 (0.95, 1.07) | 0.90 (0.82, 0.98) | 1.04 (0.98, 1.10) |
| Other (pre-2011) | 0.85 (0.77, 0.95) | 0.93 (0.85, 1.02) | 0.92 (0.78, 1.09) | 1.02 (0.91, 1.15) |
| Other (post-2011) | 0.89 (0.78, 1.02) | 0.95 (0.84, 1.08) | 0.92 (0.77, 1.11) | 0.94 (0.82, 1.09) |
| **AoU** |  |  |  |  |
| All | 0.81 (0.81, 0.82) | 0.82 (0.82, 0.83) | 0.85 (0.85, 0.86) | 0.87 (0.86, 0.87) |
| All-Race-adjusted | 0.86 (0.85, 0.86) | 0.88 (0.87, 0.88) | 0.88 (0.87, 0.88) | 0.89 (0.89, 0.90) |
| NHW | 0.81 (0.80, 0.82) | 0.80 (0.79, 0.81) | 0.88 (0.87, 0.88) | 0.89 (0.89, 0.90) |
| NHB | 0.96 (0.94, 0.97) | 0.98 (0.97, 0.99) | 0.90 (0.88, 0.92) | 0.90 (0.88, 0.92) |
| Hispanic | 0.86 (0.85, 0.87) | 0.90 (0.89, 0.91) | 0.85 (0.83, 0.86) | 0.89 (0.88, 0.9) |
| NHA | 0.87 (0.81, 0.93) | 0.89 (0.84, 0.94) | 0.96 (0.93, 0.99) | 0.97 (0.95, 0.99) |
| Multiracial | 0.85 (0.80, 0.91) | 0.83 (0.79, 0.87) | 0.90 (0.85, 0.94) | 0.91 (0.88, 0.94) |
| Other | 0.94 (0.85, 1.04) | 0.79 (0.73, 0.86) | 0.92 (0.86, 0.98) | 0.92 (0.87, 0.96) |
| None of these | 0.85 (0.79, 0.91) | 0.86 (0.81, 0.91) | 0.86 (0.82, 0.91) | 0.89 (0.85, 0.93) |

**Supplemental Table 5: Model diagnostics: P values from modified, Satterthwaite-adjusted likelihood ratio tests (NHANES) and likelihood ratio tests (AoU) testing whether addition of race-by-SES interactions to fully adjusted models improve model fit.**

| Model outcome | Model SES exposure | NHANES | AoU |
| --- | --- | --- | --- |
| T2D | Educational Attainment | p = 0.023 | p <0.001 |
| T2D | Income | p = 0.314 | p <0.001 |
| Obesity | Educational Attainment | p <0.001 | p <0.001 |
| Obesity | Income | p = 0.024 | p <0.001 |

**Supplemental Figure 1: Flow diagram of participant exclusion from the NHANES analysis.**

| **NHANES participants 2011-2018  (N = 101,316)** | |  |
| --- | --- | --- |
|  |  | **Excluded due to:**   - **Age < 18 (N = 42,105)** |
| **Adult NHANES participants 2011-2018  (N = 59,211)** | |  |
|  |  | **Excluded due to:**   - **Missing demographic information (N=7)** - **Missing both educational attainment and income-to-poverty ratio (N=865)** - **Missing all elements used to define outcome definitions (N=1)** |
| **Adult NHANES participants with complete exposure and outcome information  (N = 58,539)** | |  |
|  |  | **Excluded due to:**   - **Missing covariates (insurance status, smoking status; N=3,547)** |
| **Adult NHANES participants with complete information (N = 54,991)** | |  |

**Supplemental Figure 2: Flow diagram of participant exclusion from the All of Us cohort analysis.**

| **AoU participants with either education or income available (N = 393,840)** | | |  |
| --- | --- | --- | --- |
|  | |  | **Excluded due to:**   - **Age < 18 (N = 0)** |
| **Adult AoU participants with either education or income available (N = 393,840)** | | |  |
|  | |  | **Excluded due to:**   - **Missing demographic information (N = 3,494)** - **Missing all elements used to define outcome definitions (N = 93,277)** |
| **Adult AoU participants with complete exposure and outcome information  (N = 304,279)** | | |  |
|  | |  | **Excluded due to:**   - **Missing covariates (insurance status, smoking status; N = 7,210)** |
| **Adult AoU participants with complete information (N = 297,069)** | | |  |
|  |  | | **Excluded due to :**   - **Equivocal form of diabetes (e.g., documented both type 1 and type 2 diabetes or met insulin criteria only; N= 21,777)** |
| **Adult AoU Sample (N = 275,292)** | | |  |

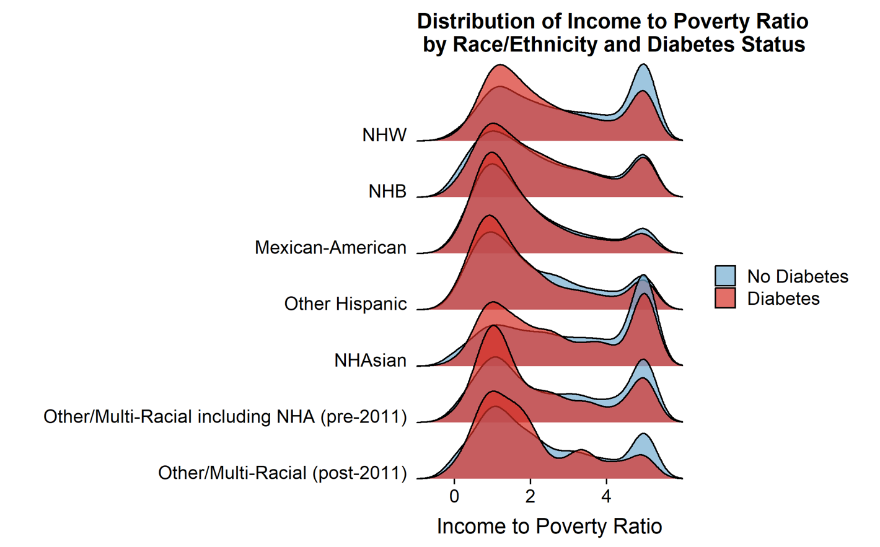

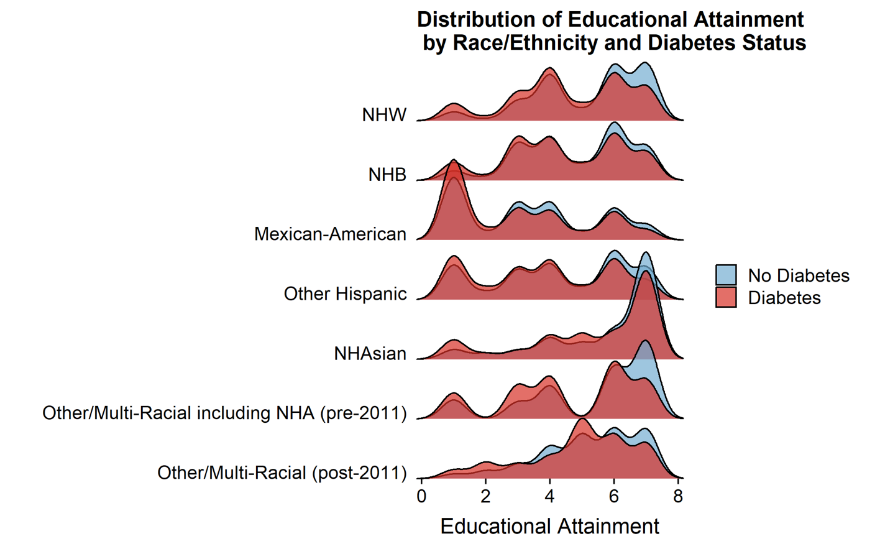
**Supplemental Figure 3: Distribution of (A,C) educational attainment and (B,D) income-to-poverty ratio within racial and ethnic groups, with and without type 2 diabetes, in the National Health and Nutrition Examination Survey 1999-2018 (A-B) and the All of Us cohort (C-D).**

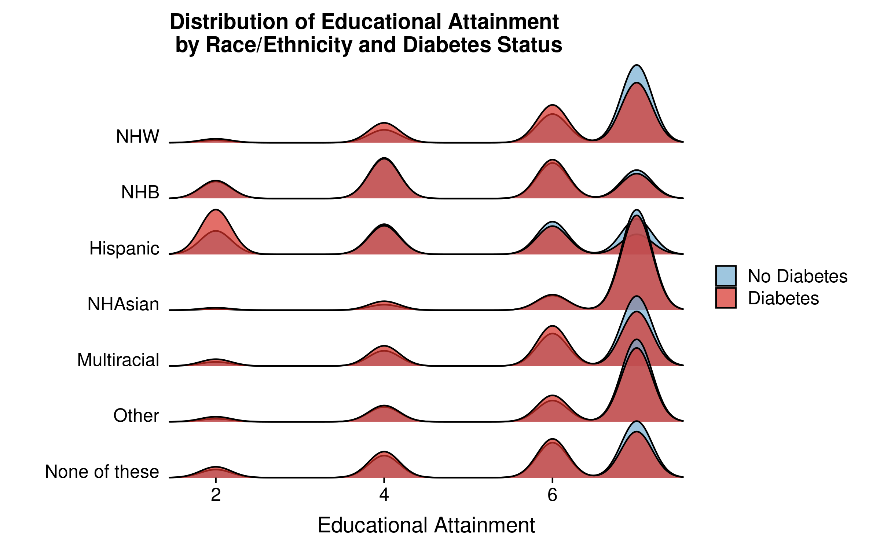

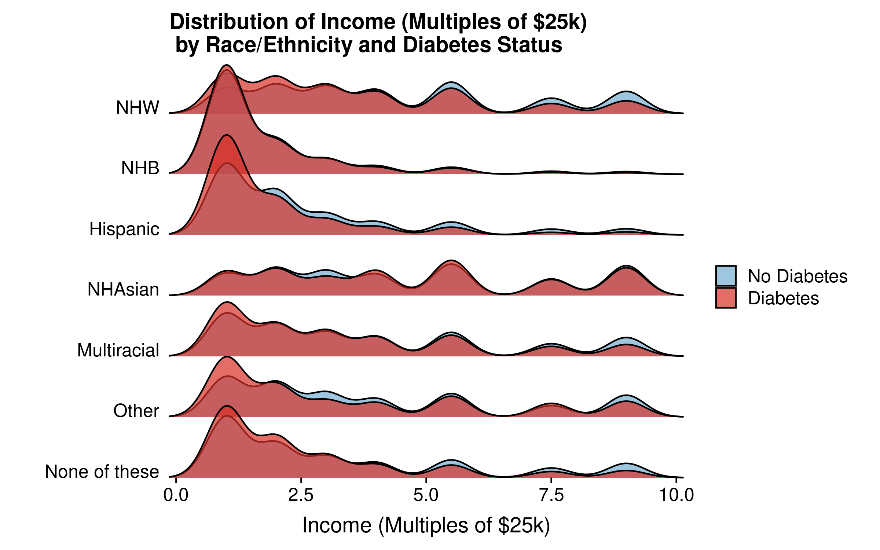

**Supplemental Figure 4: Expansion of Figure 2, including underrepresented racial and ethnic groups: Forest plots of the adjusted association of educational attainment, modeled continuously (left column) or categorically (right column), with (A) type 2 diabetes and (B) obesity prevalence, in the population overall and in each racial and ethnic subgroup, in NHANES (circles) and AoU (triangles), including racial and ethnic groups less well-represented in the data sets.**

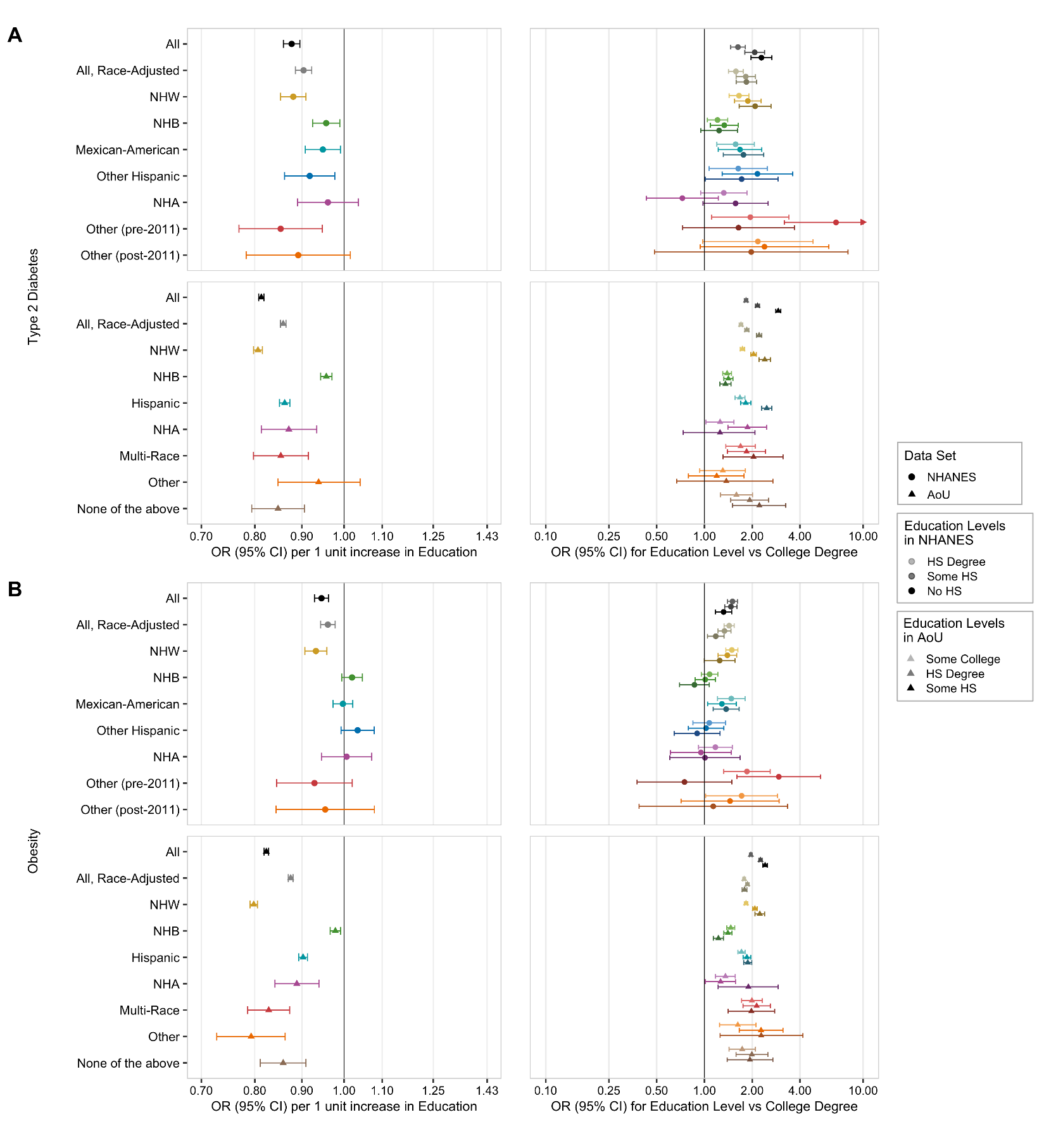

**Supplemental Figure 5: Expansion of Figure 3, including underrepresented racial and ethnic groups: Forest plots of the adjusted association of income, modeled continuously (left column) or categorically (right column), with (A) type 2 diabetes and (B) obesity prevalence, in the population overall and in each racial and ethnic subgroup, in NHANES (circles) and AoU (triangles), including racial and ethnic groups less well-represented in the data sets.**

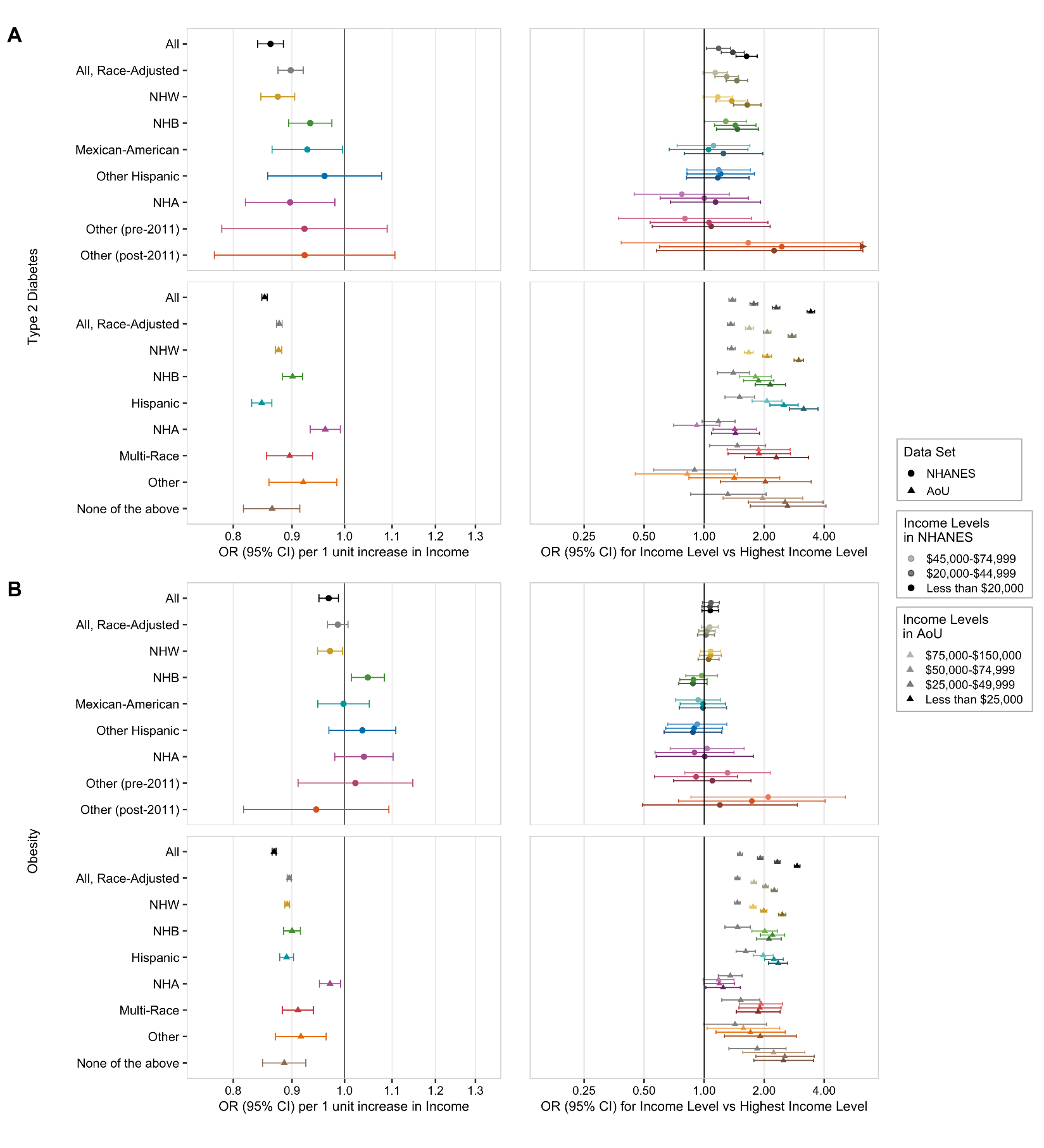

**Supplemental Figure 6: Sensitivity analysis: Forest plots of the adjusted association of continuous educational attainment with (A) type 2 diabetes and (B) obesity prevalence, in the population overall and in each racial and ethnic subgroup, in NHANES and AoU, using alternative modeling parameters (Quasi-binomial regression (left column), modified Poisson regression (middle column), and Poisson regression (right column).**

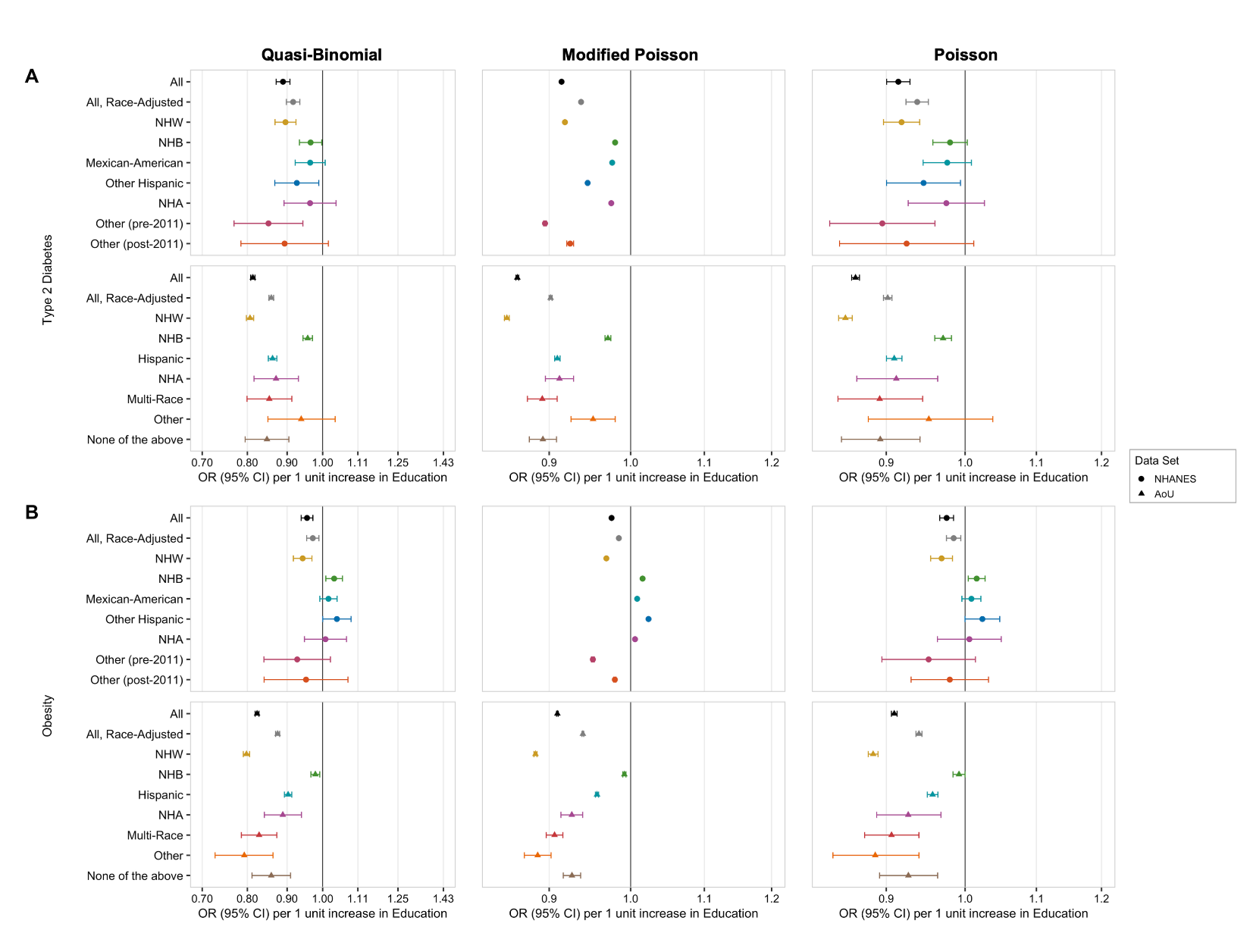

**Supplemental Figure 7: Sensitivity analysis: Forest plots of the adjusted association of continuous educational attainment with (A) type 2 diabetes and (B) obesity prevalence, in the population overall and in each racial and ethnic subgroup, in NHANES and AoU, using alternative modeling parameters (Quasi-binomial regression (left column), modified Poisson regression (middle column), and Poisson regression (right column).**

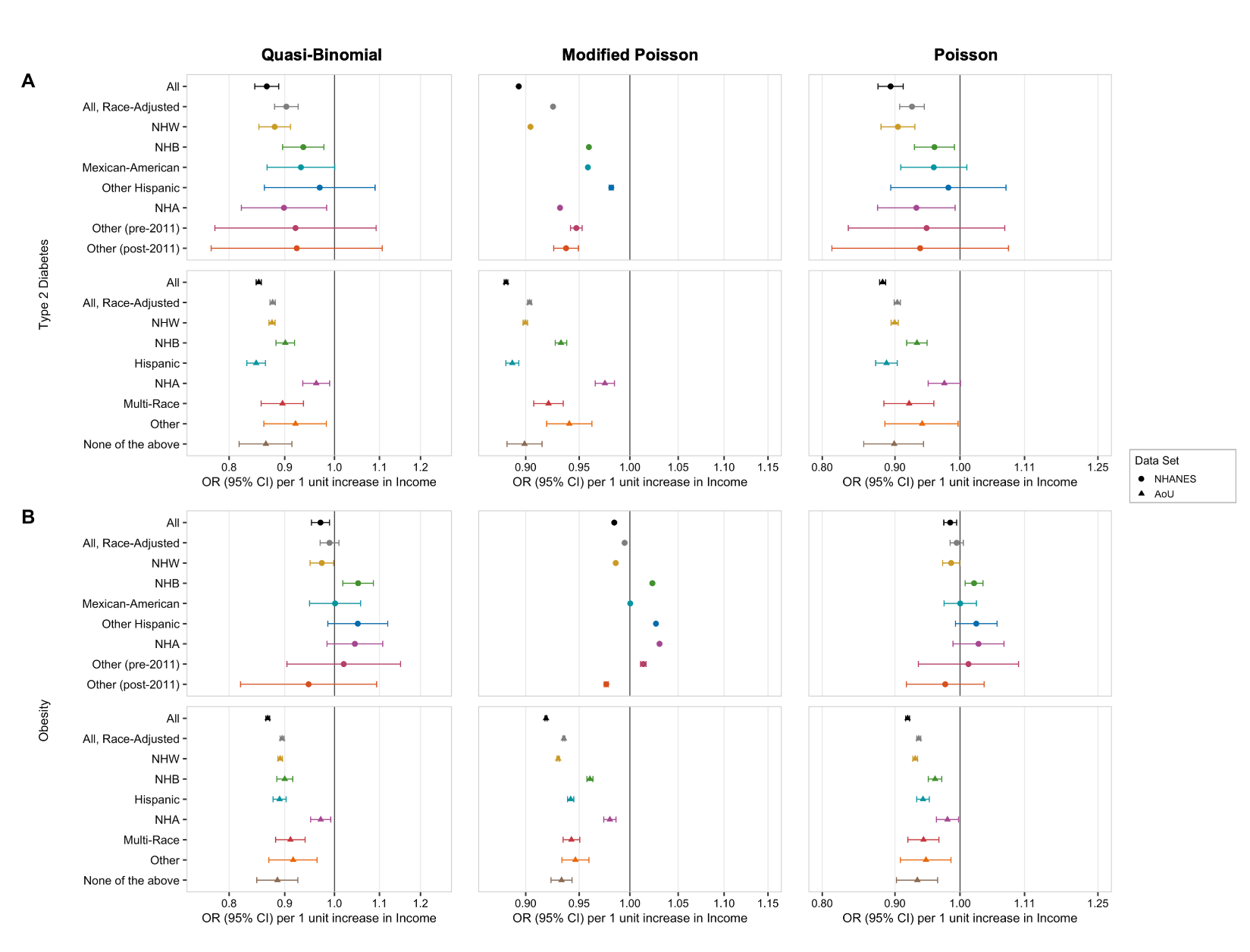
